# Genomic Epidemiology of 2023-2024 Oropouche Outbreak in Iquitos, Peru reveals independent origin from a concurrent outbreak in Brazil

**DOI:** 10.1101/2024.12.08.24318674

**Authors:** Maribel Paredes Olortegui, Francesca Schiaffino, Pablo Peñataro_Yori, Josh M. Colston, Valentino Shapiama_Lopez, Tackeshy Pinedo_Vasquez, Paul F. Garcia_Bardales, Thomas G. Flynn, Cesar Ramal-Asayag, Holley R Hughes, Emily Davis, Brandy J. Russell, Aaron C. Brault, Yuri Alfonso Alegre Palomino, Cesar Munayco, Jie Liu, Eric Houpt, Kerry K. Cooper, Craig T. Parker, Margaret N. Kosek

**Affiliations:** Asociación Benéfica Prisma, Iquitos, Peru; Division of Infectious Diseases and International Health, School of Medicine, University of Virginia, VA, USA; Faculty of Veterinary Medicine, Universidad Peruana Cayetano Heredia, San Martin de Porres, Lima, Peru; School of Human Medicine, Universidad Nacional de la Amazonia Peruana, Iquitos, Peru; Research Laboratory of Infectious and Tropical Diseases, Hospital Regional de Loreto “Felipe Santiago Arriola Iglesias”, Iquitos, Peru; Arboviral Diseases Branch, Division of Vector-borne Diseases, US Center for Disease Control and Prevention, Fort Collins, CO, USA; Gerencia Regional de Salud, Gobierno Regional de Loreto, Loreto, Peru; Centro Nacional de Epidemiologia, Prevención y Control de Enfermedades, Ministerio de Salud, Peru; School of Public Health, Qingdao University, Qingdao, China; School of Animal and Comparative Biomedical Sciences, University of Arizona, Tucson, AZ; Agricultural Research Service, U.S. Department of Agriculture, Produce Safety and Microbiology Research Unit, Albany, CA, USA

**Keywords:** arbovirus, orthobunyavirus, acute febrile illness

## Abstract

Oropouche virus is an arbovirus endemic to the Americas. Periodic outbreaks have occurred since its description in 1955. In late 2023, an outbreak occurred in Peru, centered in and around Iquitos in the Eastern Peruvian Amazon. An existing acute febrile illness (AFI) surveillance program was able to document its emergence and characterize arthralgia and dysuria and the absence of diarrhea as distinctive clinical features of Oropouche virus-associated febrile illness relative to other causes of AFI. Sequencing of isolates from the outbreak demonstrated that strains from this region were distinct from those causing disease in Brazil, despite the large-scale movement of people along the Amazon corridor, but highly similar to strains from Colombia and Ecuador. Our findings suggest that the current outbreak in South America is fundamentally multifocal in origin and not the result of geographic spread from Brazil, which experienced an outbreak between 2022 and 2024.

## Introduction

Oropouche virus (OROV) is one of the most important vector-borne diseases in Latin America, with half a million estimated cases in South America, Central America, and the Caribbean since its initial description in 1955 (1–3). Although periodic outbreaks have been well documented since then, the extent of each outbreak is poorly defined mainly because of the limited availability of diagnostics for a disease that is not clearly clinically differentiated from other etiologies of acute febrile illness (AFI). However, OROV infection and transmission have been documented clearly in the region since the late 1990’s (4–7).

The Oropouche species complex comprises orthobunyaviruses in the Simbu serogroup with three genomic segments (L, M and S). These segments frequently undergo reassortment, leading to related but distinct viruses, such as Iquitos, Madre de Dios and Perdões viruses (5, 8–10). Other human pathogenic orthobunyaviruses have been identified in the region of Loreto (such as the Itaya virus and Bellavista virus)(11, 12), yet their overall burden, distribution, and epidemiology remain obscure. The most thoroughly implicated vector in the urban cycle is *Culicoides parensis* goeldi (13), a midge species with a range extending from subtropical South America to well within the territory of the Southern United States. There is some evidence that the *Culex quinquefasciatus* mosquito is also involved in urban OROV transmission cycles, but vectorial capacity is much lower (14–16). The sylvatic cycle of OROV and its role in the spillover of orthobunyaviruses into the urban cycle of disease transmission remains understudied but has been proposed to include *Aedes serratus* and *Cuquillettidia venezuelensis* as spillover vectors (17, 18). OROV has been detected in non-human neotropical primates, sloths, and rodents, and a variety of wild birds have been found to be seropositive (19).

Clinically, Oropouche manifests itself as other arboviral etiologies of AFI. The most common symptoms include fever, myalgia, and back pain. However, meningitis and encephalitis have also been reported (20, 21). Risk factors for OROV disease have been poorly characterized. Still, several groups have noted that outbreaks tend to occur in the rainy season and areas of recent deforestation or forest fragmentation (22). In prior epidemics, dispersion patterns are thought to result from human movement among or to urban localities with the potential urban vector *C. parensis* is found (23–25).

OROV has been on the rise in Latin America and the Caribbean in 2024 (26), with cases identified in Colombia and Brazil (27), as well as in travelers from Cuba (28). The Pan American Health Organization (PAHO, World Health Organization) issued an alert on February 2, 2024 (29). The current study presents the clinical, epidemiological, and genomic findings of an outbreak of OROV in Iquitos, Loreto, Peru, from December 1^st^, 2023 to August 31^st^, 2024 as part of the ongoing RIVERA case-control study of AFI (30) and uses available regional sequences to trace the origin of the outbreak strain.

## Methods

### Study Design and Participant Enrollment

The present study is nested within the RIVERA study, a prospective health facility-based case-control study of acute febrile illness initiated in 2020 and is still ongoing in Iquitos, Peru. The study design, enrollment, and diagnostic details of the RIVERA study have been described previously (30). Briefly, patients aged ten years or older seeking care for acute febrile illness (cases) at selected facilities in Iquitos, Loreto, Peru, were enrolled, as well as age- and site-matched controls with no AFI symptoms. Both cases and controls undergo a baseline clinical assessment, and contribute blood and nasopharyngeal samples, which are tested for a locally relevant panel of pathogens by PCR. In a subsequent household visit 3-4 weeks later, additional epidemiological information is collected. Fifty case-control dyads are enrolled each month, to ensure statistical power to detect a five-fold change in the monthly prevalence of a given pathogen with a baseline prevalence of 1%, assuming 80% power and a 95% confidence level. The detection rate of approximately 1% is conservative and derived from other regional studies showing this is the interepidemic prevalence of emerging infectious diseases such as Oropouche and Mayaro virus (31).

### Sample and Data Collection at Baseline and Early Convalescence

At enrollment, whole blood and mid-turbinate samples were obtained from each case and control participant, as well as clinical and epidemiologic information. Clinical information at baseline included the presence or absence of pre-existing diseases, the presence or absence of clinical signs and symptoms in the prior two weeks, main vital signs (temperature (° C), heart rate (beats per minute), respiratory rate (breaths per minute), oxygen saturation (%), systolic and diastolic blood pressure (mmHg)) and anthropometric measurements (weight (kg), height (cm)). Demographic and epidemiologic information included age (10-19 years, 20-39 years, 40-59 years, >= 60 years), sex (female or male), area of residence (“rural areas,” defined as those participants living in Mazan or other areas outside the Iquitos metropolitan area, vs. “urban areas,” defined as participants residing in the Iquitos metropolitan area), travel in the past 15 days (no travel, travel by river, travel by other mode), the presence or absence of ectoparasites, bats or rodents on the participant’s body or at home.

Participant follow-up was completed 28 days after enrollment. Field workers contacted cases and controls using instant messaging and visited their households to ascertain their health status. A whole blood and mid-turbinate sample were obtained, as were follow-up clinical information and additional epidemiological data. Follow-up clinical data included the same clinical signs and symptoms collected at baseline. Sociodemographic information collected included labor and educational information from the head of the household, household characteristics, and the presence or absence of animals in the house.

### Statistical Analysis

This nested case-control analysis aimed to characterize both the clinical and the risk factor profile of OROV infection in the RIVERA study. Different definitions of cases and controls were used from the parent study and for each of these two analysis components. For the analysis of clinical OROV disease symptomology, cases were defined as OROV-positive AFI cases and controls as AFI cases that were negative for all pathogens tested (unattributed AFI). For the analysis of risk factors for OROV infection, by contrast, cases were defined as any OROV-positive subjects (whether symptomatic or not), and controls as asymptomatic subjects (RIVERA controls) that were negative for all pathogens tested. For both parts of the analysis, cases were matched with up to 5 controls each by broad age group (10 – 24 years, 25 – 49 years, 50+ years), sex, area of residence (Iquitos Metropolitan Area or Mazan and other rural areas), and whether they were enrolled before or since the start of the OROV outbreak (December 1^st^, 2023). Odds ratios and their 95% confidence intervals (95% CI) for OROV positivity were calculated for each of the categorical clinical and epidemiological variables previously described using conditional logistic regression to account for matching using fixed effects within matched case-control sets. For continuous variables, equivalent fixed effects linear models were fitted to compare mean values

### Sample Processing and OROV Diagnosis

Total nucleic acids (TNA) were extracted from whole blood samples using the High Pure Viral Nucleic Acid Large Volume Kit (Roche Life Science, Indianapolis, IN) as instructed by manufacturers. TNA from whole blood samples was tested for OROV using TaqMan array cards (Thermo Fisher Scientific, Waltham, MA). The pathogens targeted by this array card are presented in **Supplementary Figure 1**. Samples with a cycle threshold (Ct) of less or equal to 35 were considered positive. The OROV primers and probe utilized included: Forward: 5’-TGATCCGGAGGCAGCATA-3’, Reverse: 5’-ACACCAGCATTGAGCACTTG-3’, Probe: FAM-CCGTATCTAGCTTCAAATGCC-MGB (32). Mid turbinate swabs were tested for Influenza A, Influenza B, and SARS-CoV-2 using the CDC Influenza SARS-CoV-2 multiplex assay as described previously (30).

### OROV Culture

Cellular fractions of EDTA anticoagulated blood with Ct values less than 30 were sent to the Arboviral Diseases Branch, Centers for Disease Control and Prevention for virus culture. Each sample was inoculated in volumes of 2ul, 20ul, and 100ul along with 2ml of DMEM (Gibco, Waltham, MA, USA) maintenance media supplemented with 2% fetal bovine serum (Seradign, Radnor, PA, USA) into confluent T25 flasks (Corning) of Vero cells and incubated for one hour at 37°C for adsorption. Eight ml of DMEM maintenance media was added to each flask and incubated at 37°C. Flasks were visualized under light microscopy daily for the presence of cytopathic effects (CPE). When CPE impacted 50-75% of the cell monolayer the supernatant was harvested and centrifuged at 10,000 rpm for 10 minutes to remove cell debris. Two 0.5 ml aliquots were removed for down stream testing and the remainder was frozen as bulk.

### OROV Culture Based Sequencing

RNA was extracted from samples using the QiaAmp Viral RNA Mini kit (Qiagen, Georgetown, MD, USA) following the manufacturer’s protocol. The sample was treated using TURBODNase using the manufacturers protocol (ThermoFisher, Waltham, MA, USA). Complementary DNA was generated using random hexamer amplification via the Ovation RNA-seq v2 kit (Tecan, Morrisville, NC, USA)) followed by library preparation using the DNAPrep kit (Illumina, San Diego, CA, USA). Libraries were quantified with the Qubit 4 fluorometer and the dsDNA High-sensitivity kit (ThermoFisher, Waltham, MA, USA). Library size was confirmed on the Tapestation 2200 (Agilent, Santa Clara, CA, USA). Libraries were pooled and loaded at a final concentration of 8 pM onto a MiSeq V2 300-cycle flow cell (Illumina, San Diego, CA, USA).

FASTQ files were trimmed of adapter sequences and low-quality reads removed (Q<30) using Illumina BCL Convert (V4.0). *De novo* assembly was completed using the rnaviral presets of the SPAdes assembler (V3.15.3). Viral contigs were identified using the BLASTn (V2.15.0) nt_viruses database and confirmed using the BLASTn nt database.

### Whole Blood Amplicon Based Sequencing

Amplicon-based sequencing was done using the TNA extract of OROV positive whole blood samples with a Ct value under 30. We adapted an amplicon-based sequencing protocol from Quick et al. (33) for Zika and did primer enrichment using the primers specified in Wise *et al*. (34) for OROV run on an Oxford Nanopore Technology (ONT) system. Genome assembly will be performed using the Map to Reference application within Geneious Prime. The ONT reads were trimmed to Q15 and then mapped to the individual *in silico* amplicon sequences to create consensus sequences for each amplicon. Finally, the consensus amplicons, which overlap, were assembled into complete genome segments (L, M, and S).

Oropouche genomes are available under NCBI Bio project **PRJNA813162**.

### Genomic Analysis

Publicly available OROV sequences for the L, M, and S segments from 2024 from Brazil, Colombia, Peru and Cuba, from 2016 from Ecuador, and from Peru prior to 2023 were obtained from NCBI Virus (35). Any sequence containing more than a single unknown (e.g., N) nucleotide was eliminated from the analysis to avoid these unknown nucleotides biasing the analysis. Ultimately, 17 strains representing various regions of Brazil, two strains from Colombia, six strains from Ecuador, one strain from Cuba, and 11 strains from Peru were chosen (**Supplemental Table 1**) and used for phylogenetic comparison against Peruvian viral sequences from 2024 generated during this study. To determine the best model to utilize for the phylogenetic analysis, each of the alignments was tested using ModelTest-NG (v0.1.7) (36) and for each segment, it was determined that the General Time Reversible (GTR) model with gamma-distributed rate variation was the most appropriate. Maximum likelihood trees for each segment were generated using this model and bootstrapping 10,000 times and then visualized and annotated using the Interactive Tree of Life (iTOL) (37) online tool. Each branch on the maximum likelihood tree was colored according to the country of isolation (Colombia – orange, Ecuador – green, Brazil – light red, Cuba – light blue, Peru 2024 - blue, and historical Peru (prior to 2024) – black). Each taxa is labeled with the country of isolation, year of isolation, and a number indicating which sample from that country it represents, which the number allows for tracking each segment of the same OROV among the three maximum likelihood trees.

Each viral segment was processed individually for the analysis, and the sequences for each segment were aligned using the MUSCLE plugin (v5.1) (38) in Geneious Prime (v2024.0.7) (39) using the Perturbed Profile-Profile (PPP) algorithm with five hidden Markov model (HMM) perturbations. Each alignment was then trimmed to make sure each sequence included the exact same number of nucleotides across identical regions to avoid bias due to extra nucleotides for some sequences. Overall, the S segment alignment was trimmed to 640 nucleotides, the M segment was trimmed to 4,088 nucleotides, and the L segment was trimmed to 6,550 nucleotides, and then each of these trimmed alignments was exported as a Phylip format alignment to generate maximum likelihood trees using RAxML(v8.2.13) (40).

### Ethics Statement

The RIVERA study has been approved by the Institutional Review Board of Asociación Benéfica Prisma (CE0855.20) (FWA00001219), University of Virginia (FWA0006183), and Hospital Regional de Loreto. Additional approval has been obtained by the Research Commission of the Regional Health Direction of Loreto. Written informed consent was obtained for all participants. For children aged 10–18, both parental written informed consent and written informed assent were obtained. All participants consented to the further use of biological specimens.

## Results

### Epidemiology and Symptoms Associated with Illness

The RIVERA study initiated in March 2021 and enrollment continues at this time. Before December 2023, OROV viremia was detected in 0.4% of RIVERA subjects (6 detections in AFI cases, 7 in asymptomatic controls).

Between December 1^st^, 2023, and August 31^st^, 2024, OROV viremia were detected in 39 whole blood samples (32 in AFI cases, 7 in asymptomatic controls) out of 834 samples processed in that same time frame (**Figure 1**). At the epidemic peak in the first three months of 2024, a detection rate of 8.7% was recorded, representing a more than 20-fold increase in OROV-positivity compared to pre-outbreak levels. OROV was therefore detectable in asymptomatic individuals before and during the epidemic. Coinfection of OROV with DENV, Histoplasma, and M. tuberculosis, was observed in one asymptomatic subject each, while two cases each of OROV/SARS-CoV-2 and OROV/influenza A coinfection were detected in AFI cases.

**Figure 1.**
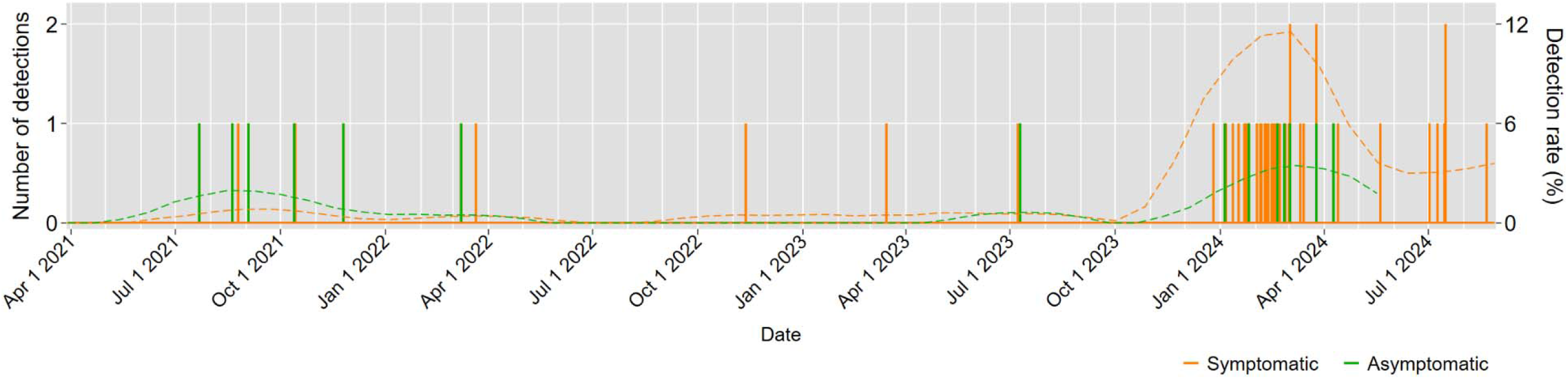
Time series graph of OROV detections in symptomatic and asymptomatic RIVERA subjects over the three year study period reveal sporadic detections of Oropouche in 0.4% of cases and control prior to December 2023. Between December 2023 and August 2024, Oropouche identification increased to a peak prevalence of 11% in those with acute febrile illness and 3% of asymptomatic controls in March 2024.

No deaths were associated with the detected acute clinical illnesses, and no cases of meningoencephalitis were observed. All cases assessed at 30 days of follow-up, had clinically recovered from their illnesses with no sequelae.

Clinical symptoms and their associations with OROV-positive AFI are reported in **Table 1**. Several such features of illness were identified that were significantly more common in those febrile with Oropouche compared to unattributed cases of acute febrile illness. These symptoms included joint pain, which 5.07 (95% CI:1.66, 15.45) times the odds of occurring, and dysuria, which was 3.63 (95% CI: 1.13,11.63) in Oropouche cases relative to those with undifferentiated febrile illness. Diarrhea was less likely to be present in those with febrile disease caused by Oropouche (OR 0.28: 95% CI 0.09, 0.85) than in individuals with unattributable febrile illness. Headache, myalgia, abdominal pain, nausea, vomiting, and rashes were not observed to be more strongly associated with Oropouche disease than an undefined febrile illness.

**Table 1:**
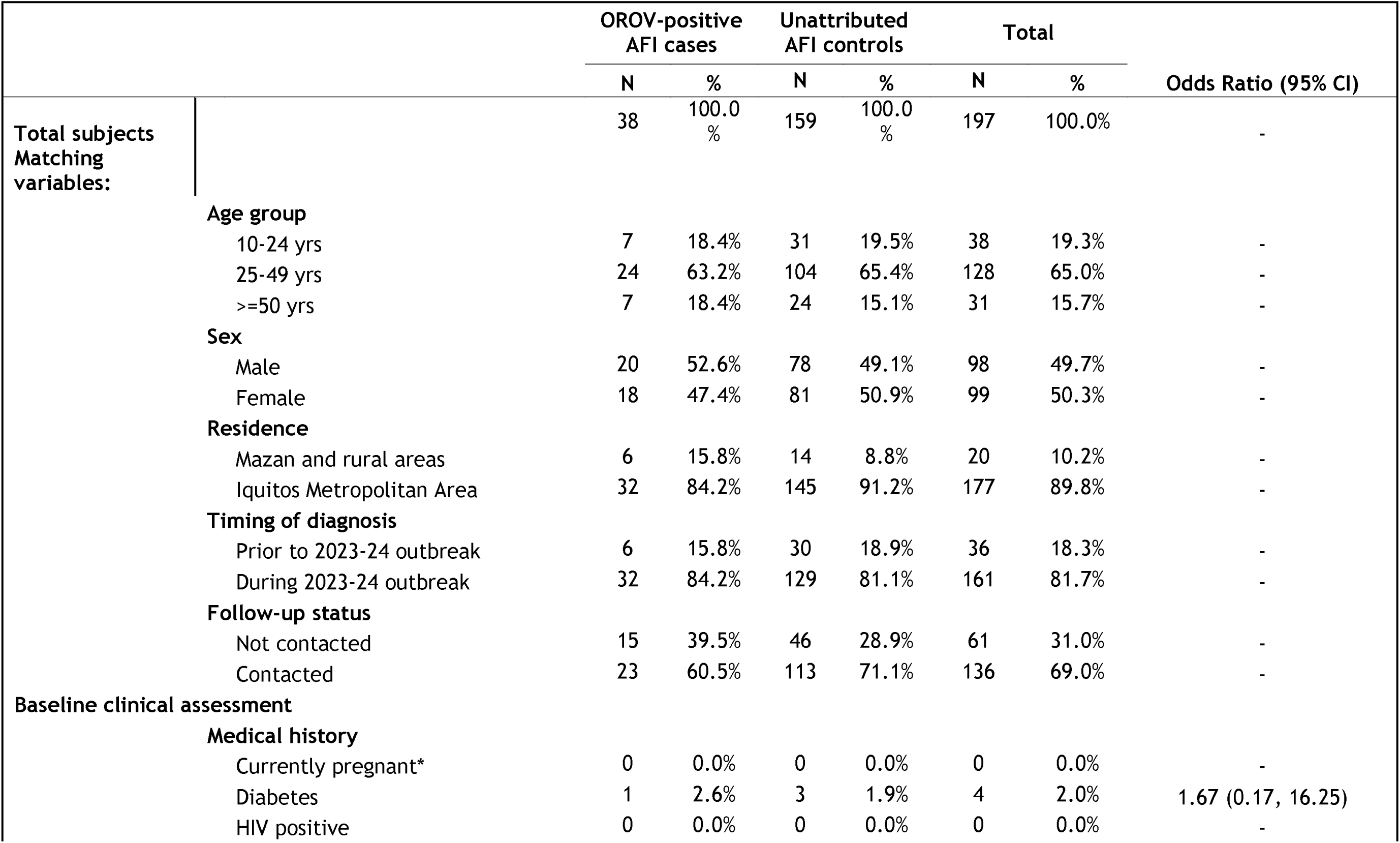
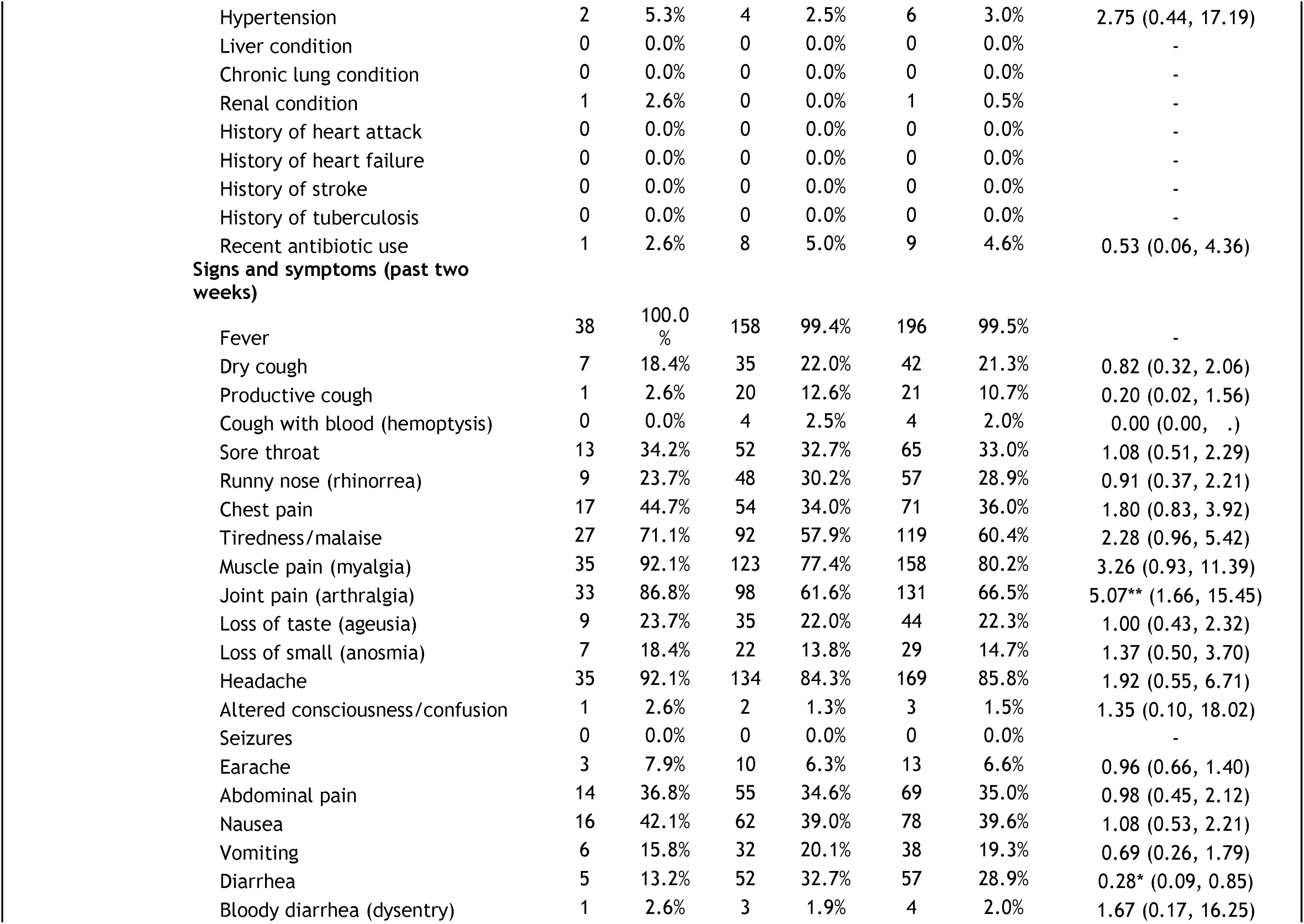
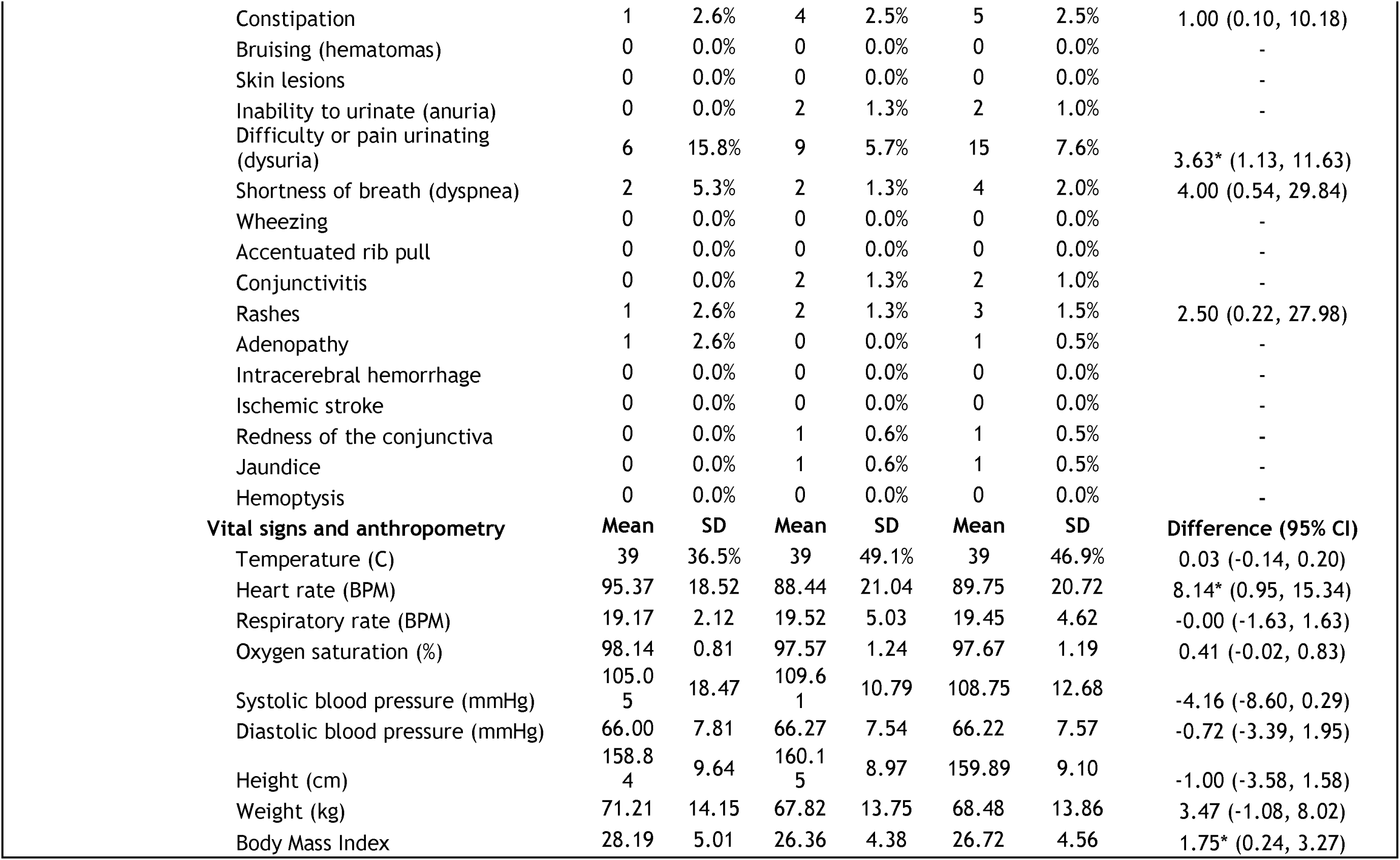
Distribution of matching variables, medical history, signs, symptoms, vital signs and anthropometry in OROV-positive AFI cases and unattributed AFI controls and the odds ratios (with 95% confidence intervals) for their associations with the OROV-positive AFI outcome.

|  |  | OROV-positive<br>AFI cases |  | Unattributed<br>AFI controls |  | Total |  | Odds Ratio (95% CI) |
| --- | --- | --- | --- | --- | --- | --- | --- | --- |
|  |  | N | % | N | % | N | % |  |
| **Total subjects** |  | 38 | 100.0<br>% | 159 | 100.0<br>% | 197 | 100.0% | - |
| **Matching variables:** |  |  |  |  |  |  |  |  |
|  | **Age group** |  |  |  |  |  |  |  |
|  | 10-24 yrs | 7 | 18.4% | 31 | 19.5% | 38 | 19.3% | - |
|  | 25-49 yrs | 24 | 63.2% | 104 | 65.4% | 128 | 65.0% | - |
|  | >=50 yrs | 7 | 18.4% | 24 | 15.1% | 31 | 15.7% | - |
|  | **Sex** |  |  |  |  |  |  |  |
|  | Male | 20 | 52.6% | 78 | 49.1% | 98 | 49.7% | - |
|  | Female | 18 | 47.4% | 81 | 50.9% | 99 | 50.3% | - |
|  | **Residence** |  |  |  |  |  |  |  |
|  | Mazan and rural areas | 6 | 15.8% | 14 | 8.8% | 20 | 10.2% | - |
|  | Iquitos Metropolitan Area | 32 | 84.2% | 145 | 91.2% | 177 | 89.8% | - |
|  | **Timing of diagnosis** |  |  |  |  |  |  |  |
|  | Prior to 2023-24 outbreak | 6 | 15.8% | 30 | 18.9% | 36 | 18.3% | - |
|  | During 2023-24 outbreak | 32 | 84.2% | 129 | 81.1% | 161 | 81.7% | - |
|  | **Follow-up status** |  |  |  |  |  |  |  |
|  | Not contacted | 15 | 39.5% | 46 | 28.9% | 61 | 31.0% | - |
|  | Contacted | 23 | 60.5% | 113 | 71.1% | 136 | 69.0% | - |
| **Baseline clinical assessment** |  |  |  |  |  |  |  |  |
|  | **Medical history** |  |  |  |  |  |  |  |
|  | Currently pregnant* | 0 | 0.0% | 0 | 0.0% | 0 | 0.0% | - |
|  | Diabetes | 1 | 2.6% | 3 | 1.9% | 4 | 2.0% | 1.67 (0.17, 16.25) |
|  | HIV positive | 0 | 0.0% | 0 | 0.0% | 0 | 0.0% | - |

|  |  |  |  |  |  |  |  |
| --- | --- | --- | --- | --- | --- | --- | --- |
| Hypertension | 2 | 5.3% | 4 | 2.5% | 6 | 3.0% | 2.75 (0.44, 17.19) |
| Liver condition | 0 | 0.0% | 0 | 0.0% | 0 | 0.0% | - |
| Chronic lung condition | 0 | 0.0% | 0 | 0.0% | 0 | 0.0% | - |
| Renal condition | 1 | 2.6% | 0 | 0.0% | 1 | 0.5% | - |
| History of heart attack | 0 | 0.0% | 0 | 0.0% | 0 | 0.0% | - |
| History of heart failure | 0 | 0.0% | 0 | 0.0% | 0 | 0.0% | - |
| History of stroke | 0 | 0.0% | 0 | 0.0% | 0 | 0.0% | - |
| History of tuberculosis | 0 | 0.0% | 0 | 0.0% | 0 | 0.0% | - |
| Recent antibiotic use | 1 | 2.6% | 8 | 5.0% | 9 | 4.6% | 0.53 (0.06, 4.36) |
| **Signs and symptoms (past two weeks)** |  |  |  |  |  |  |  |
| Fever | 38 | 100.0% | 158 | 99.4% | 196 | 99.5% | - |
| Dry cough | 7 | 18.4% | 35 | 22.0% | 42 | 21.3% | 0.82 (0.32, 2.06) |
| Productive cough | 1 | 2.6% | 20 | 12.6% | 21 | 10.7% | 0.20 (0.02, 1.56) |
| Cough with blood (hemoptysis) | 0 | 0.0% | 4 | 2.5% | 4 | 2.0% | 0.00 (0.00, .) |
| Sore throat | 13 | 34.2% | 52 | 32.7% | 65 | 33.0% | 1.08 (0.51, 2.29) |
| Runny nose (rhinorrhea) | 9 | 23.7% | 48 | 30.2% | 57 | 28.9% | 0.91 (0.37, 2.21) |
| Chest pain | 17 | 44.7% | 54 | 34.0% | 71 | 36.0% | 1.80 (0.83, 3.92) |
| Tiredness/malaise | 27 | 71.1% | 92 | 57.9% | 119 | 60.4% | 2.28 (0.96, 5.42) |
| Muscle pain (myalgia) | 35 | 92.1% | 123 | 77.4% | 158 | 80.2% | 3.26 (0.93, 11.39) |
| Joint pain (arthralgia) | 33 | 86.8% | 98 | 61.6% | 131 | 66.5% | 5.07** (1.66, 15.45) |
| Loss of taste (ageusia) | 9 | 23.7% | 35 | 22.0% | 44 | 22.3% | 1.00 (0.43, 2.32) |
| Loss of smell (anosmia) | 7 | 18.4% | 22 | 13.8% | 29 | 14.7% | 1.37 (0.50, 3.70) |
| Headache | 35 | 92.1% | 134 | 84.3% | 169 | 85.8% | 1.92 (0.55, 6.71) |
| Altered consciousness/confusion | 1 | 2.6% | 2 | 1.3% | 3 | 1.5% | 1.35 (0.10, 18.02) |
| Seizures | 0 | 0.0% | 0 | 0.0% | 0 | 0.0% | - |
| Earache | 3 | 7.9% | 10 | 6.3% | 13 | 6.6% | 0.96 (0.66, 1.40) |
| Abdominal pain | 14 | 36.8% | 55 | 34.6% | 69 | 35.0% | 0.98 (0.45, 2.12) |
| Nausea | 16 | 42.1% | 62 | 39.0% | 78 | 39.6% | 1.08 (0.53, 2.21) |
| Vomiting | 6 | 15.8% | 32 | 20.1% | 38 | 19.3% | 0.69 (0.26, 1.79) |
| Diarrhea | 5 | 13.2% | 52 | 32.7% | 57 | 28.9% | 0.28* (0.09, 0.85) |
| Bloody diarrhea (dysentery) | 1 | 2.6% | 3 | 1.9% | 4 | 2.0% | 1.67 (0.17, 16.25) |
| Constipation | 1 | 2.6% | 4 | 2.5% | 5 | 2.5% | 1.00 (0.10, 10.18) |
| Bruising (hematomas) | 0 | 0.0% | 0 | 0.0% | 0 | 0.0% | - |
| Skin lesions | 0 | 0.0% | 0 | 0.0% | 0 | 0.0% | - |
| Inability to urinate (anuria) | 0 | 0.0% | 2 | 1.3% | 2 | 1.0% | - |
| Difficulty or pain urinating (dysuria) | 6 | 15.8% | 9 | 5.7% | 15 | 7.6% | 3.63* (1.13, 11.63) |
| Shortness of breath (dyspnea) | 2 | 5.3% | 2 | 1.3% | 4 | 2.0% | 4.00 (0.54, 29.84) |
| Wheezing | 0 | 0.0% | 0 | 0.0% | 0 | 0.0% | - |
| Accentuated rib pull | 0 | 0.0% | 0 | 0.0% | 0 | 0.0% | - |
| Conjunctivitis | 0 | 0.0% | 2 | 1.3% | 2 | 1.0% | - |
| Rashes | 1 | 2.6% | 2 | 1.3% | 3 | 1.5% | 2.50 (0.22, 27.98) |
| Adenopathy | 1 | 2.6% | 0 | 0.0% | 1 | 0.5% | - |
| Intracerebral hemorrhage | 0 | 0.0% | 0 | 0.0% | 0 | 0.0% | - |
| Ischemic stroke | 0 | 0.0% | 0 | 0.0% | 0 | 0.0% | - |
| Redness of the conjunctiva | 0 | 0.0% | 1 | 0.6% | 1 | 0.5% | - |
| Jaundice | 0 | 0.0% | 1 | 0.6% | 1 | 0.5% | - |
| Hemoptysis | 0 | 0.0% | 0 | 0.0% | 0 | 0.0% | - |
| **Vital signs and anthropometry** | **Mean** | **SD** | **Mean** | **SD** | **Mean** | **SD** | **Difference (95% CI)** |
| Temperature (C) | 39 | 36.5% | 39 | 49.1% | 39 | 46.9% | 0.03 (-0.14, 0.20) |
| Heart rate (BPM) | 95.37 | 18.52 | 88.44 | 21.04 | 89.75 | 20.72 | 8.14* (0.95, 15.34) |
| Respiratory rate (BPM) | 19.17 | 2.12 | 19.52 | 5.03 | 19.45 | 4.62 | -0.00 (-1.63, 1.63) |
| Oxygen saturation (%) | 98.14 | 0.81 | 97.57 | 1.24 | 97.67 | 1.19 | 0.41 (-0.02, 0.83) |
| Systolic blood pressure (mmHg) | 105.05 | 18.47 | 109.61 | 10.79 | 108.75 | 12.68 | -4.16 (-8.60, 0.29) |
| Diastolic blood pressure (mmHg) | 66.00 | 7.81 | 66.27 | 7.54 | 66.22 | 7.57 | -0.72 (-3.39, 1.95) |
| Height (cm) | 158.84 | 9.64 | 160.15 | 8.97 | 159.89 | 9.10 | -1.00 (-3.58, 1.58) |
| Weight (kg) | 71.21 | 14.15 | 67.82 | 13.75 | 68.48 | 13.86 | 3.47 (-1.08, 8.02) |
| Body Mass Index | 28.19 | 5.01 | 26.36 | 4.38 | 26.72 | 4.56 | 1.75* (0.24, 3.27) |

Epidemiological risk factors and their associations with OROV infection are reported in **Table 2**. A history of travel in the preceding 15 days was associated with a 4.46 increase in the odds of OROV detection compared with pathogen- and symptom-negative controls, although the majority of OROV infection cases (84.6%) reported no travel. Individuals in which OROV was detected had a BMI that was 1.8 kg/m^2^ higher than pathogen and symptom negative controls.

**Table 2:**
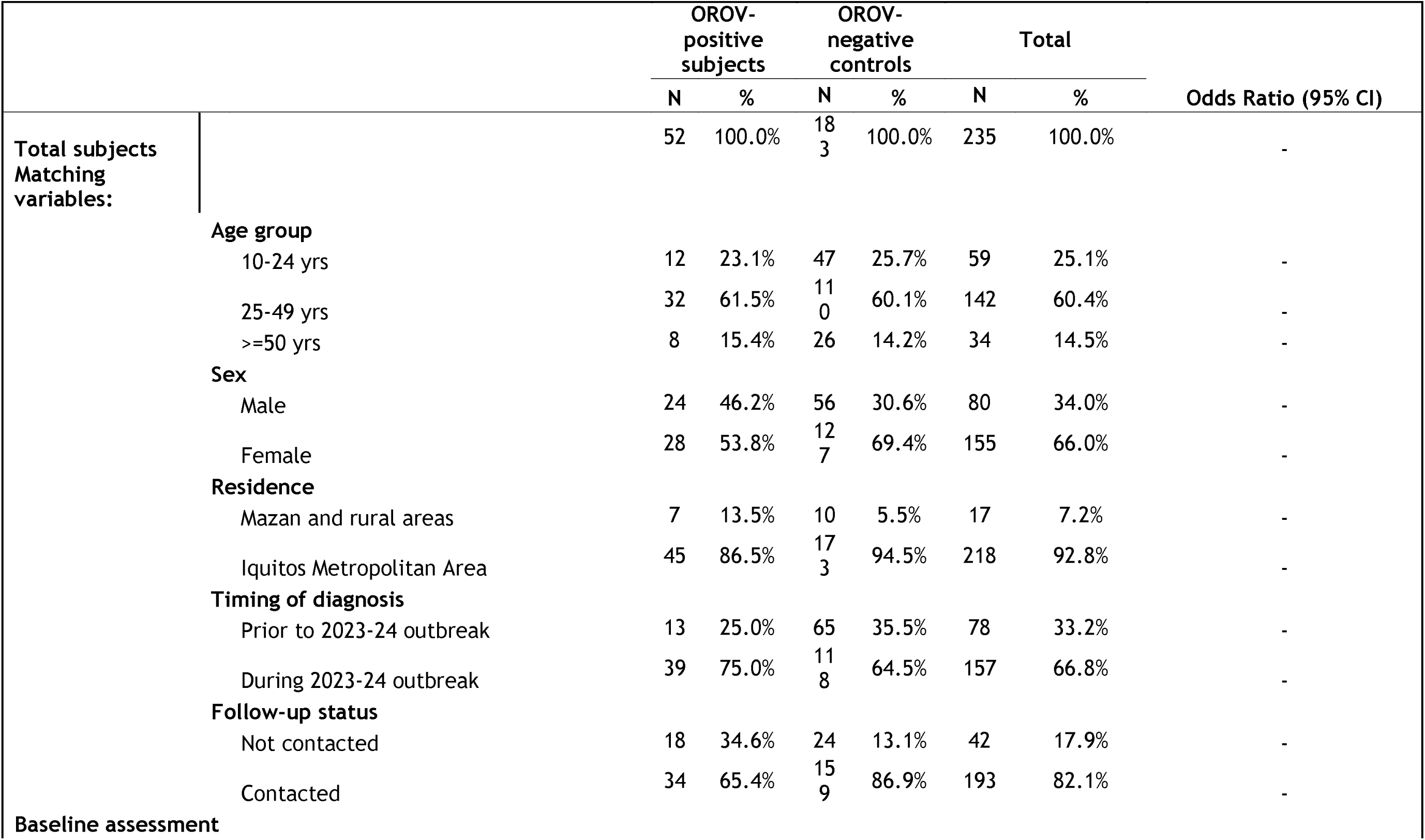
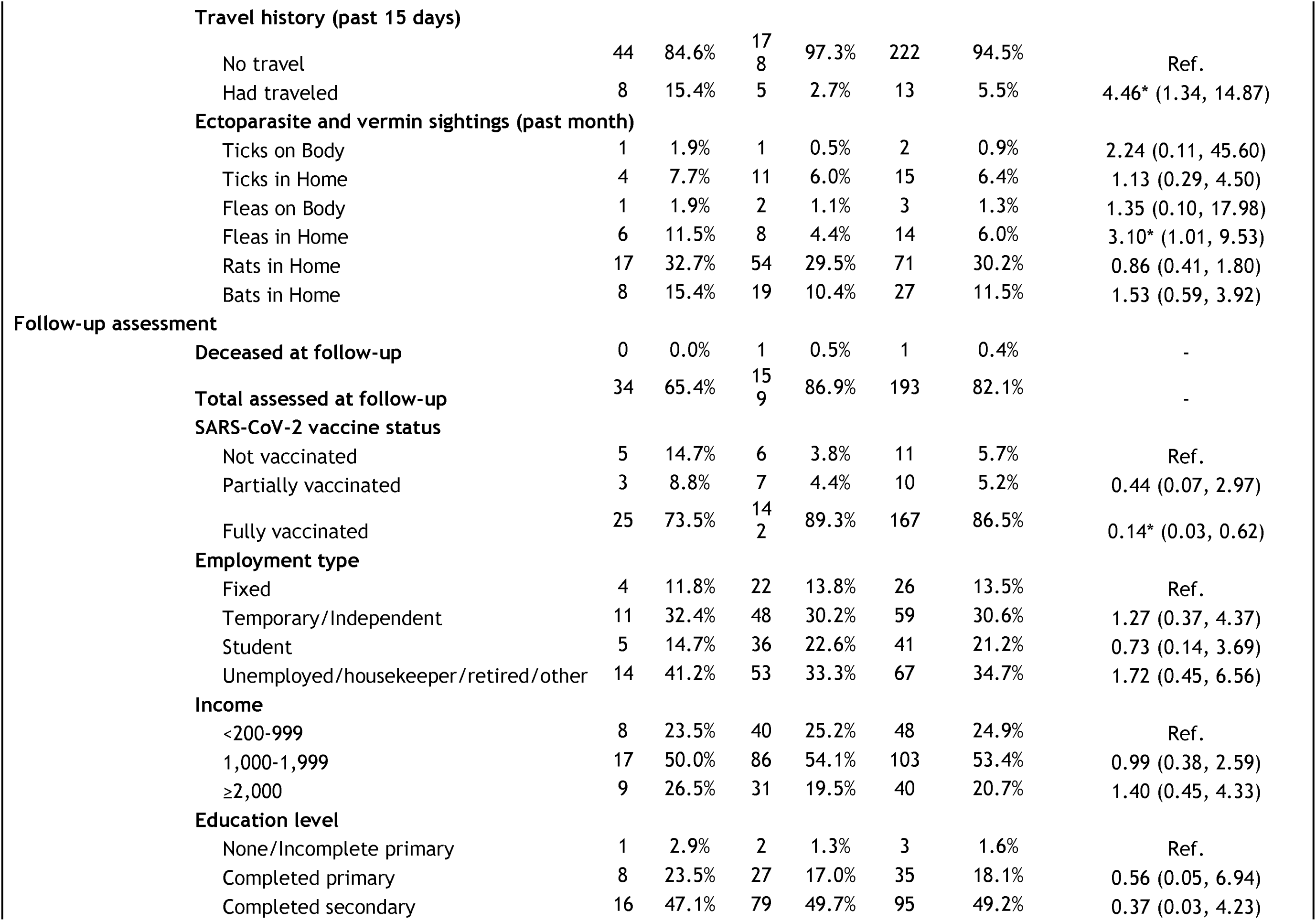
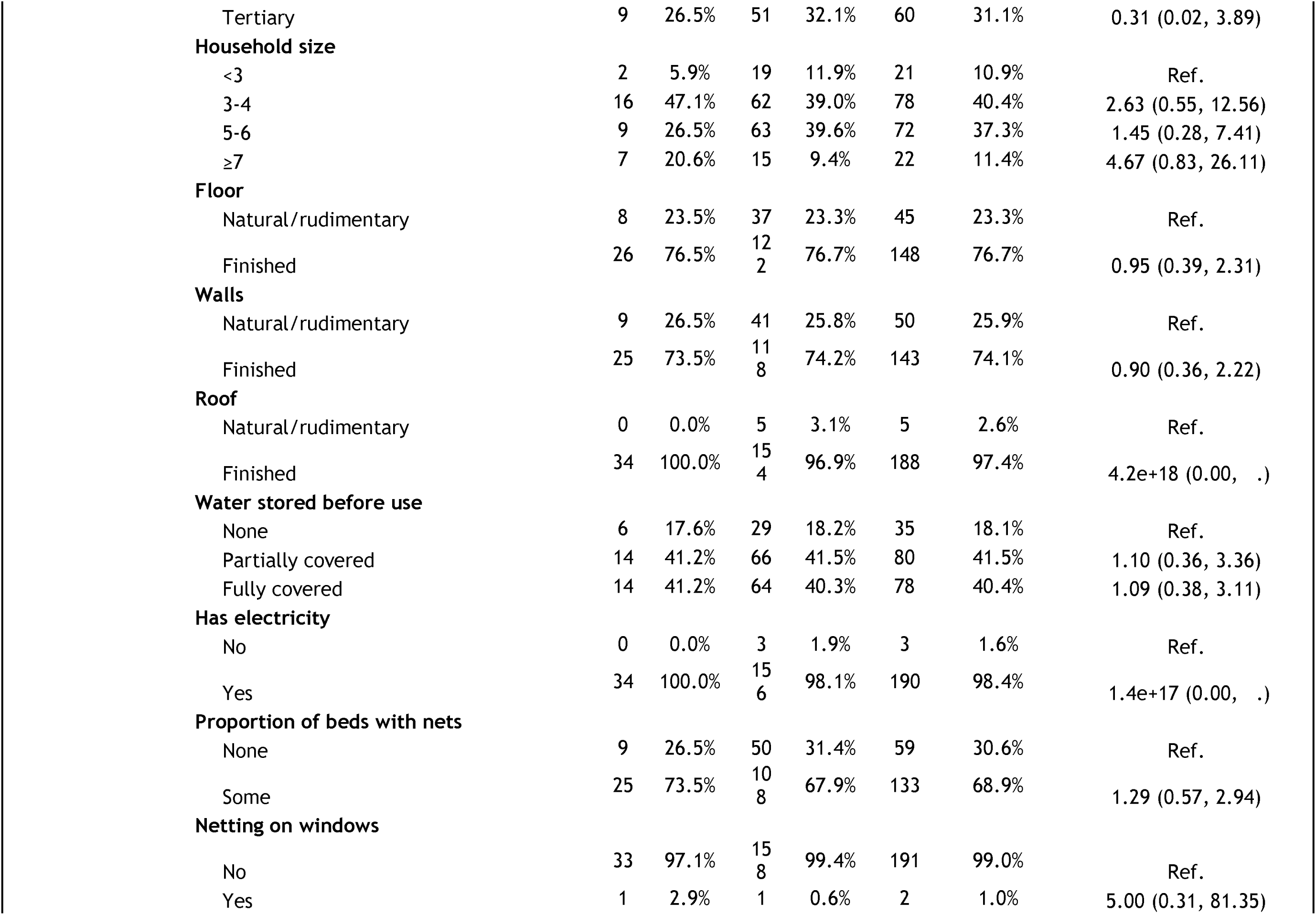
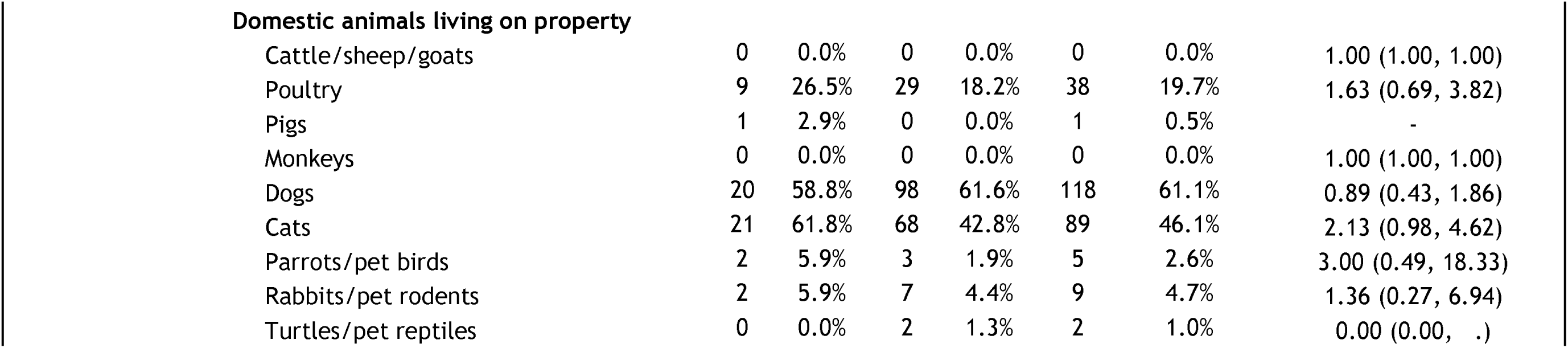
Distribution of matching variables and epidemiological risk factors ascertained at baseline assessment and at 4-week follow-up in OROV-positive subjects and pathogen-negative asymptomatic controls.

|  |  | OROV-positive<br>subjects |  | OROV-negative<br>controls |  | Total |  | Odds Ratio (95% CI) |
| --- | --- | --- | --- | --- | --- | --- | --- | --- |
|  |  | N | % | N | % | N | % |  |
| Total subjects<br>Matching<br>variables: |  | 52 | 100.0% | 183 | 100.0% | 235 | 100.0% | - |
|  | **Age group** |  |  |  |  |  |  |  |
|  | 10-24 yrs | 12 | 23.1% | 47 | 25.7% | 59 | 25.1% | - |
|  | 25-49 yrs | 32 | 61.5% | 110 | 60.1% | 142 | 60.4% | - |
|  | >=50 yrs | 8 | 15.4% | 26 | 14.2% | 34 | 14.5% | - |
|  | **Sex** |  |  |  |  |  |  |  |
|  | Male | 24 | 46.2% | 56 | 30.6% | 80 | 34.0% | - |
|  | Female | 28 | 53.8% | 127 | 69.4% | 155 | 66.0% | - |
|  | **Residence** |  |  |  |  |  |  |  |
|  | Mazan and rural areas | 7 | 13.5% | 10 | 5.5% | 17 | 7.2% | - |
|  | Iquitos Metropolitan Area | 45 | 86.5% | 173 | 94.5% | 218 | 92.8% | - |
|  | **Timing of diagnosis** |  |  |  |  |  |  |  |
|  | Prior to 2023-24 outbreak | 13 | 25.0% | 65 | 35.5% | 78 | 33.2% | - |
|  | During 2023-24 outbreak | 39 | 75.0% | 118 | 64.5% | 157 | 66.8% | - |
|  | **Follow-up status** |  |  |  |  |  |  |  |
|  | Not contacted | 18 | 34.6% | 24 | 13.1% | 42 | 17.9% | - |
|  | Contacted | 34 | 65.4% | 159 | 86.9% | 193 | 82.1% | - |
| Baseline assessment |  |  |  |  |  |  |  |  |

|  |  |  |  |  |  |  |  |
| --- | --- | --- | --- | --- | --- | --- | --- |
| **Travel history (past 15 days)** |  |  |  |  |  |  |  |
| No travel | 44 | 84.6% | 17<br>8 | 97.3% | 222 | 94.5% | Ref. |
| Had traveled | 8 | 15.4% | 5 | 2.7% | 13 | 5.5% | 4.46* (1.34, 14.87) |
| **Ectoparasite and vermin sightings (past month)** |  |  |  |  |  |  |  |
| Ticks on Body | 1 | 1.9% | 1 | 0.5% | 2 | 0.9% | 2.24 (0.11, 45.60) |
| Ticks in Home | 4 | 7.7% | 11 | 6.0% | 15 | 6.4% | 1.13 (0.29, 4.50) |
| Fleas on Body | 1 | 1.9% | 2 | 1.1% | 3 | 1.3% | 1.35 (0.10, 17.98) |
| Fleas in Home | 6 | 11.5% | 8 | 4.4% | 14 | 6.0% | 3.10* (1.01, 9.53) |
| Rats in Home | 17 | 32.7% | 54 | 29.5% | 71 | 30.2% | 0.86 (0.41, 1.80) |
| Bats in Home | 8 | 15.4% | 19 | 10.4% | 27 | 11.5% | 1.53 (0.59, 3.92) |
| **Follow-up assessment** |  |  |  |  |  |  |  |
| Deceased at follow-up | 0 | 0.0% | 1 | 0.5% | 1 | 0.4% | - |
| Total assessed at follow-up | 34 | 65.4% | 15<br>9 | 86.9% | 193 | 82.1% | - |
| **SARS-CoV-2 vaccine status** |  |  |  |  |  |  |  |
| Not vaccinated | 5 | 14.7% | 6 | 3.8% | 11 | 5.7% | Ref. |
| Partially vaccinated | 3 | 8.8% | 7 | 4.4% | 10 | 5.2% | 0.44 (0.07, 2.97) |
| Fully vaccinated | 25 | 73.5% | 14<br>2 | 89.3% | 167 | 86.5% | 0.14* (0.03, 0.62) |
| **Employment type** |  |  |  |  |  |  |  |
| Fixed | 4 | 11.8% | 22 | 13.8% | 26 | 13.5% | Ref. |
| Temporary/Independent | 11 | 32.4% | 48 | 30.2% | 59 | 30.6% | 1.27 (0.37, 4.37) |
| Student | 5 | 14.7% | 36 | 22.6% | 41 | 21.2% | 0.73 (0.14, 3.69) |
| Unemployed/housekeeper/retired/other | 14 | 41.2% | 53 | 33.3% | 67 | 34.7% | 1.72 (0.45, 6.56) |
| **Income** |  |  |  |  |  |  |  |
| <200-999 | 8 | 23.5% | 40 | 25.2% | 48 | 24.9% | Ref. |
| 1,000-1,999 | 17 | 50.0% | 86 | 54.1% | 103 | 53.4% | 0.99 (0.38, 2.59) |
| ≥2,000 | 9 | 26.5% | 31 | 19.5% | 40 | 20.7% | 1.40 (0.45, 4.33) |
| **Education level** |  |  |  |  |  |  |  |
| None/Incomplete primary | 1 | 2.9% | 2 | 1.3% | 3 | 1.6% | Ref. |
| Completed primary | 8 | 23.5% | 27 | 17.0% | 35 | 18.1% | 0.56 (0.05, 6.94) |
| Completed secondary | 16 | 47.1% | 79 | 49.7% | 95 | 49.2% | 0.37 (0.03, 4.23) |
| Tertiary | 9 | 26.5% | 51 | 32.1% | 60 | 31.1% | 0.31 (0.02, 3.89) |
| **Household size** |  |  |  |  |  |  |  |
| <3 | 2 | 5.9% | 19 | 11.9% | 21 | 10.9% | Ref. |
| 3-4 | 16 | 47.1% | 62 | 39.0% | 78 | 40.4% | 2.63 (0.55, 12.56) |
| 5-6 | 9 | 26.5% | 63 | 39.6% | 72 | 37.3% | 1.45 (0.28, 7.41) |
| ≥7 | 7 | 20.6% | 15 | 9.4% | 22 | 11.4% | 4.67 (0.83, 26.11) |
| **Floor** |  |  |  |  |  |  |  |
| Natural/rudimentary | 8 | 23.5% | 37 | 23.3% | 45 | 23.3% | Ref. |
| Finished | 26 | 76.5% | 122 | 76.7% | 148 | 76.7% | 0.95 (0.39, 2.31) |
| **Walls** |  |  |  |  |  |  |  |
| Natural/rudimentary | 9 | 26.5% | 41 | 25.8% | 50 | 25.9% | Ref. |
| Finished | 25 | 73.5% | 118 | 74.2% | 143 | 74.1% | 0.90 (0.36, 2.22) |
| **Roof** |  |  |  |  |  |  |  |
| Natural/rudimentary | 0 | 0.0% | 5 | 3.1% | 5 | 2.6% | Ref. |
| Finished | 34 | 100.0% | 154 | 96.9% | 188 | 97.4% | 4.2e+18 (0.00, .) |
| **Water stored before use** |  |  |  |  |  |  |  |
| None | 6 | 17.6% | 29 | 18.2% | 35 | 18.1% | Ref. |
| Partially covered | 14 | 41.2% | 66 | 41.5% | 80 | 41.5% | 1.10 (0.36, 3.36) |
| Fully covered | 14 | 41.2% | 64 | 40.3% | 78 | 40.4% | 1.09 (0.38, 3.11) |
| **Has electricity** |  |  |  |  |  |  |  |
| No | 0 | 0.0% | 3 | 1.9% | 3 | 1.6% | Ref. |
| Yes | 34 | 100.0% | 156 | 98.1% | 190 | 98.4% | 1.4e+17 (0.00, .) |
| **Proportion of beds with nets** |  |  |  |  |  |  |  |
| None | 9 | 26.5% | 50 | 31.4% | 59 | 30.6% | Ref. |
| Some | 25 | 73.5% | 108 | 67.9% | 133 | 68.9% | 1.29 (0.57, 2.94) |
| **Netting on windows** |  |  |  |  |  |  |  |
| No | 33 | 97.1% | 158 | 99.4% | 191 | 99.0% | Ref. |
| Yes | 1 | 2.9% | 1 | 0.6% | 2 | 1.0% | 5.00 (0.31, 81.35) |

**Domestic animals living on property**
|  |  |  |  |  |  |  |  |
| --- | --- | --- | --- | --- | --- | --- | --- |
| Cattle/sheep/goats | 0 | 0.0% | 0 | 0.0% | 0 | 0.0% | 1.00 (1.00, 1.00) |
| Poultry | 9 | 26.5% | 29 | 18.2% | 38 | 19.7% | 1.63 (0.69, 3.82) |
| Pigs | 1 | 2.9% | 0 | 0.0% | 1 | 0.5% | - |
| Monkeys | 0 | 0.0% | 0 | 0.0% | 0 | 0.0% | 1.00 (1.00, 1.00) |
| Dogs | 20 | 58.8% | 98 | 61.6% | 118 | 61.1% | 0.89 (0.43, 1.86) |
| Cats | 21 | 61.8% | 68 | 42.8% | 89 | 46.1% | 2.13 (0.98, 4.62) |
| Parrots/pet birds | 2 | 5.9% | 3 | 1.9% | 5 | 2.6% | 3.00 (0.49, 18.33) |
| Rabbits/pet rodents | 2 | 5.9% | 7 | 4.4% | 9 | 4.7% | 1.36 (0.27, 6.94) |
| Turtles/pet reptiles | 0 | 0.0% | 2 | 1.3% | 2 | 1.0% | 0.00 (0.00, .) |

### Viral culture

Four specimens with Ct of < 30 were sent to the CDC for culture. Three of the 4 samples yielded a cytopathic virus confirmed as OROV by sequencing. Cytopathic effect was first observed in the three samples two days post inoculation (DPI), by day 3 DPI flasks were ready to harvest.

### Sequencing and Genomic Analysis of OROV strains

Sequencing was done to confirm qPCR diagnostics and to reassemble complete genomes to study the origin of the outbreak strain. Full genomes were assembled from 10 distinct participants. In additional cases where full genomes were not available, sequencing confirmed the diagnosis in all examined cases.

Phylogenetic analysis was performed separately for the S, M, and L segments. Unrooted maximum likelihood trees were constructed, showing similar patterns for all segments (**Figure 2**). The sequenced 2023-2024 Iquitos isolates were highly homogeneous among the 10 sequenced strains. They were closely related to strains sequenced in Colombia in 2024 and more distantly related to strains from Ecuador in 2016, followed by strains from Peru between 1995-2008. The remaining strains analyzed for comparison were from the Brazil outbreak or clustered with this outbreak. The first was a strain from Southern Peru in the Madre de Dios region bordering on Acre (indicated as Peru NAMRU 2024 #5). The other strain to cluster in this set of isolates is a strain from a traveler returning to Italy from Cuba.

**Figure 2.**
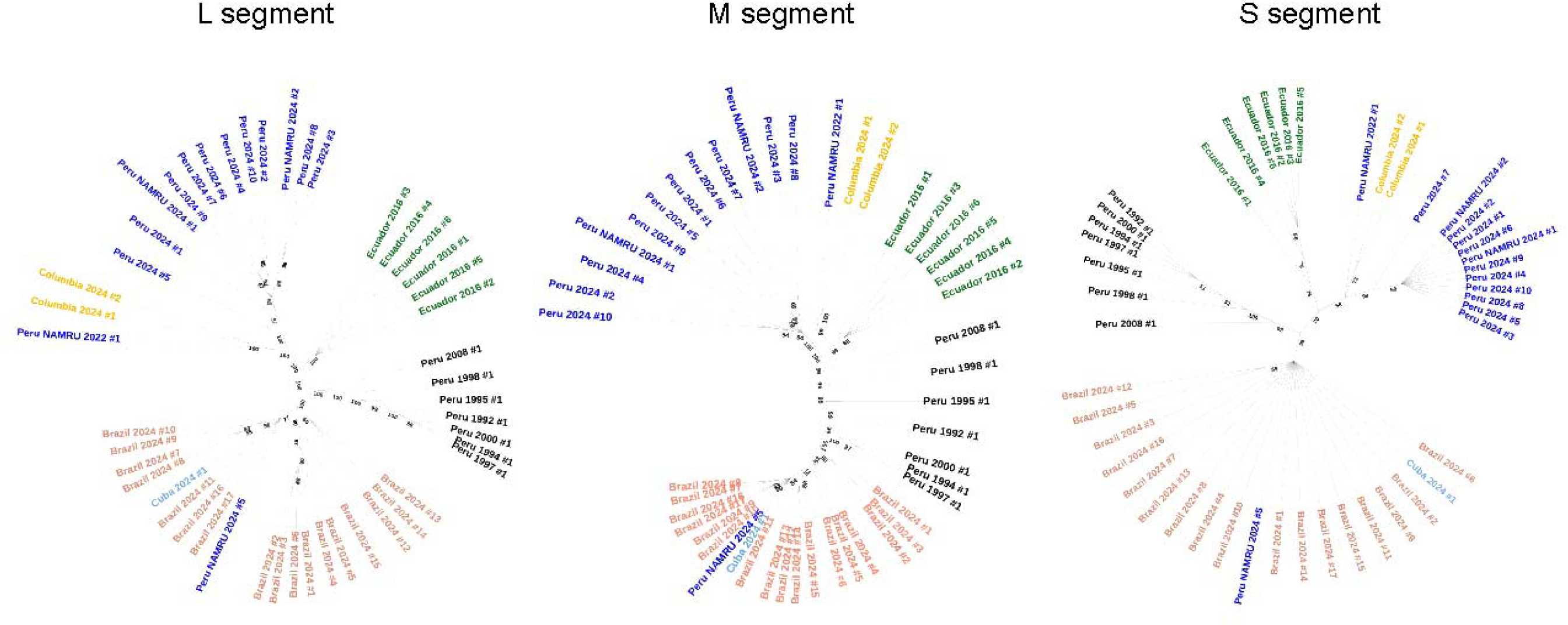
Unrooted Maximum-likelihood (ML) tree based on the (1) 6,550 bp of the L segment; (2) 4,087 bp of the M segment; and (3) 638 bp of the S segment. Nucleotide alignments for each segment were generated using MUSCLE plugin (v5.1) in Geneious Prime (v2024.0.7). Each tree was generated using the General Time Reversible (GTR) model with gamma distributed rates and bootstrapped 10,000 times.

To examine the genomic regions exhibiting differences between different geographic regions, amino acid alignment was done for each of the three genomic segments (**Figure 3**). Minimal polymorphisms were identified in the S and L segments, while polymorphisms were concentrated in the N terminus of the Gc protein on the M segment.

**Figure 3.**
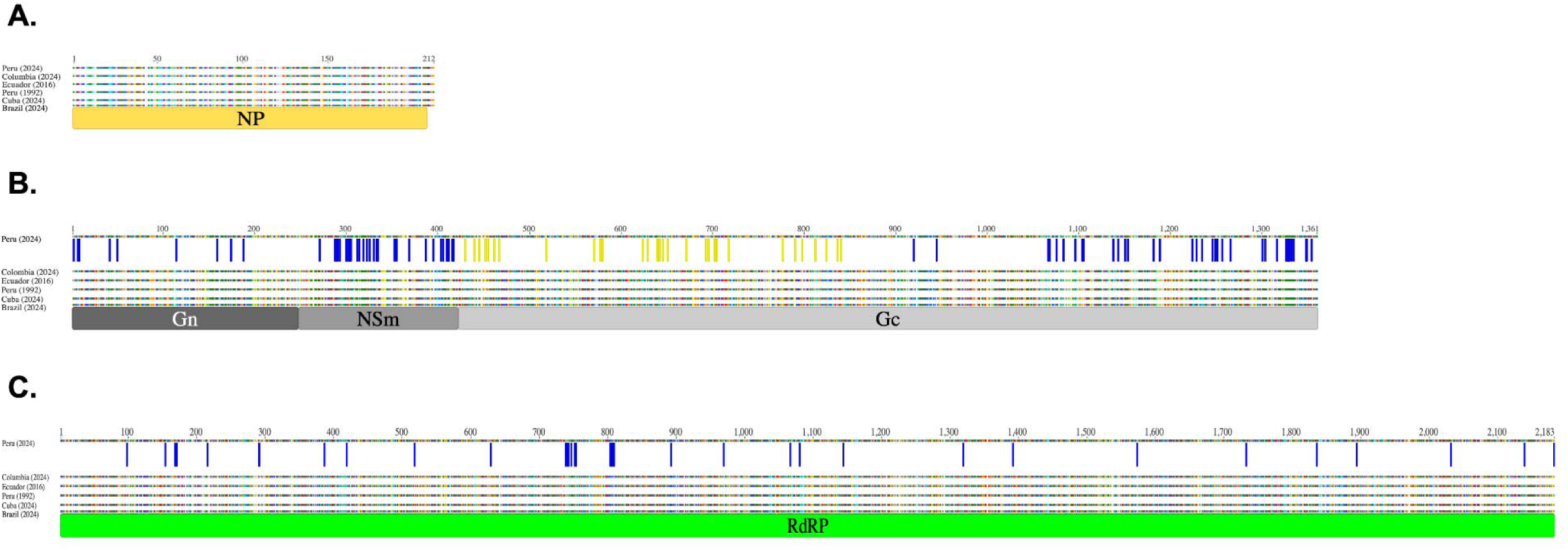
Amino acid sequence alignment for the S (A), M (B), and L (C) segments of the Oropouche virus from Peru (2024), Colombia (2024), Ecuador (2016), Peru (1992), Cuba (2024), and Brazil (2024). The S segment has 0/212 (0%) amino acid heterogeneity, and the L segment 38/2,183 (1.74%) amino acid heterogeneity among the viruses from different countries. However, there is quite a significant amount of amino acid heterogeneity in the M segment, as it has 105/1,361 (7.71%) amino acid heterogeneity, including 30/428 (7.01%) amino acid changes in the N terminus of Gc, the region coding the antigen that is purported to mediate protective immunity.

## Discussion

The RIVERA study demonstrated the ability of our health facility-based surveillance to rapidly detect an ongoing outbreak and inform genomic epidemiology in response to an outbreak of OROV in the region. Surveillance was calibrated to sample 100 samples a month, an intensity that would allow for the detection of a fivefold change in the monthly prevalence of a pathogen with a baseline prevalence of 1%, assuming an 80% power and 95% confidence level. The case-control component allowed us to delineate asymptomatic transmission. The use of a broadly diagnostic array card for targeted diagnostics of the principal pathogens causing AFI— or pathogens of significance due to the availability of vaccines or specific programmatic interventions for their treatment and control—enabled the identification of an appropriate comparator population. This allowed differentiation of symptoms in individuals with febrile illness caused by OROV from those that were negative for the primary pathogens responsible for AFI in the region. Despite the large concurrent outbreak in Brazil, our sequencing revealed that the circulating OROV strains in our study were more closely related to strains from Colombia (2020) and Ecuador (2016), both of which are thought to have originated in Peru. The N terminus of the Gc protein is the coding region for the antigen that appears to drive protective immunity based on the work of Hellert (41) and Gutierrez (42). Alignment and comparisons of the amino acids of the M segment reveal a 7% divergence in amino acids in the variable regions of the Gc protein that represents the head of the spike protein between the 2023 Brazil strains and the strains we identified from Peru in late 2023 and early 2024. The findings are of importance as they suggest that environmental factors have favored the transmission of OROV across the Amazon, but that the epidemic in Peru has a distinct ancestry and does not represent a geographic extension of OROV from Brazil, despite daily barges traveling along the Amazon river between Manaus and Iquitos and a predicted multitude of introductions from human communities traveling in each direction. Given the multitude of opportunities for the introduction of the Brazilian strains in Peru and during this two-year outbreak, it seems plausible to believe that neutralizing antibodies exist in Peru to the Brazil strain. The epidemiology of OROV is understudied, but annual seroconversion rates in the region have previously documented annual seroconversion rates of 28% in rural communities within 10 kilometers of Iquitos in the absence of an outbreak, strongly suggesting that transmission in the area is both intense and continuous (7).

Risk factors associated with an increase risk of infection were travel outside of the peri-urban zones of Iquitos and having a higher BMI.

Newly emerging pathogenic orthobunyaviruses have been detected in the Amazonian region and have been associated with genome segment reassortment (8, 11, 43–45). Such is the case of Iquitos virus, which is a product of the reassortment of the S and L segments of OROV and the M segment of an unknown Simbu serogroup virus (5). Other human pathogenic orthobunyaviruses have been identified in the region of Loreto (Itaya virus and Bellavista virus),(11, 12) yet their overall burden, distribution, and epidemiology still need to be established. The sylvatic cycle of OROV, and its role in the spillover of orthobunyaviruses into the urban cycle of disease transmission remain understudied. OROV has been detected in non-human neotropical primates, sloths, rodents, and a variety of wild birds (19). Although there is an apparently higher prevalence of OROV in avian hosts, the role of these animals in disease transmission to humans remains poorly understood. Specifically, it is critically important to understand the role of arthropods and other species of insects in the transmission of OROV from birds to humans. Evidence of larger wild mammalian species in OROV transmission is negligible. Additionally, the potential identification of OROV in domestic chickens and ducks, previously demonstrated in an outbreak investigation in Brazil (19), would open the possibility of a domestic animal reservoir within urban areas where small-scale poultry rearing is widely practiced. Therefore, establishing a human cohort and concurrently sampling arthropods, mosquitoes, avian and mammalian species will provide a unique evidence base to characterize the transmission dynamics of OROV.

## Data Availability

All genomic data is publicly available under accession numbers referenced in the manuscript.

## Supplementary Figures and Tables

**Supplemental Figure S1.**
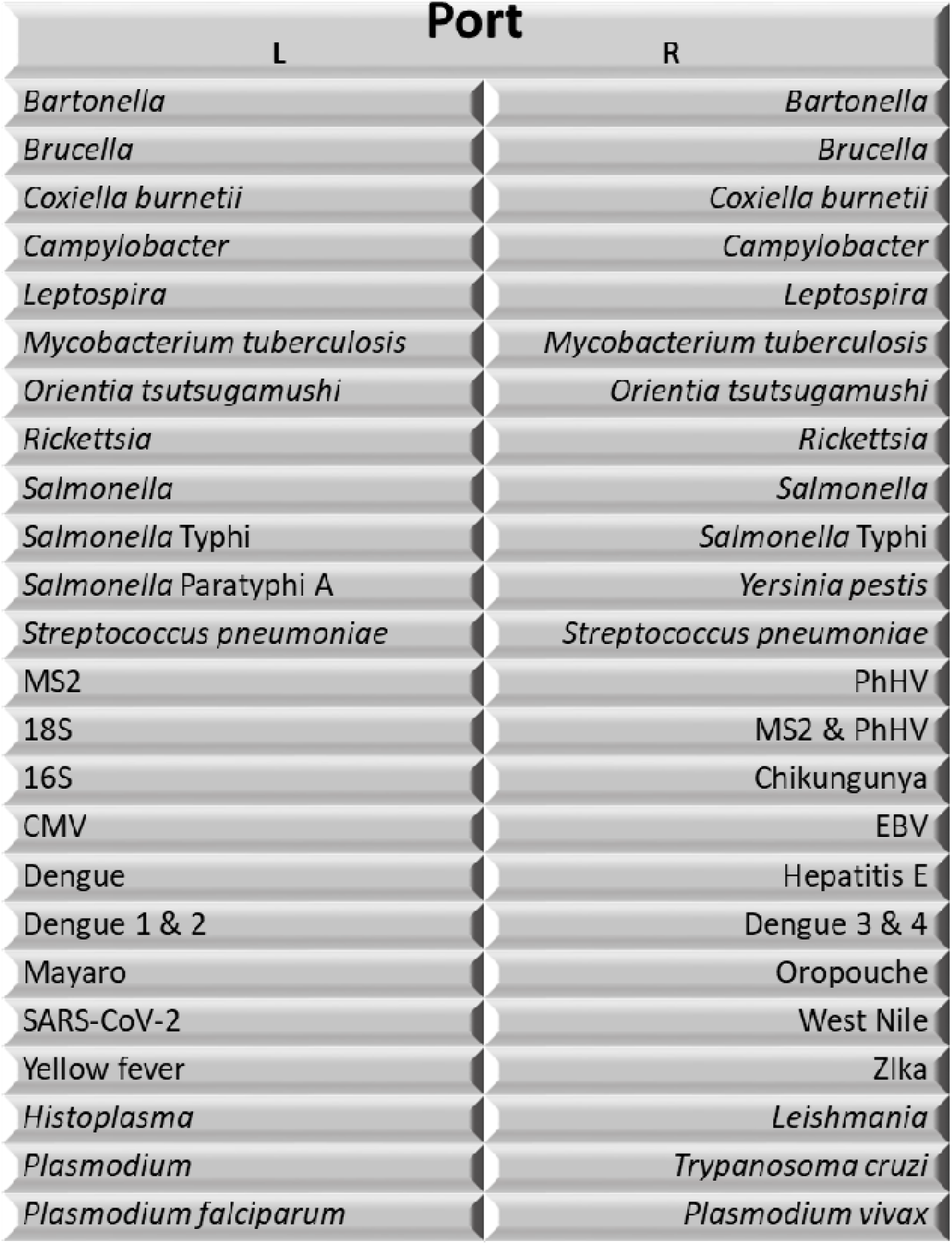
Customized Diagnostic Array Card layout for concurrent diagnosis of endemic, emerging, and vaccine preventable infectious diseases. All participants (cases and controls) had a single blood sample analyzed with this diagnostic panel.

**Supplementary Table 1.**
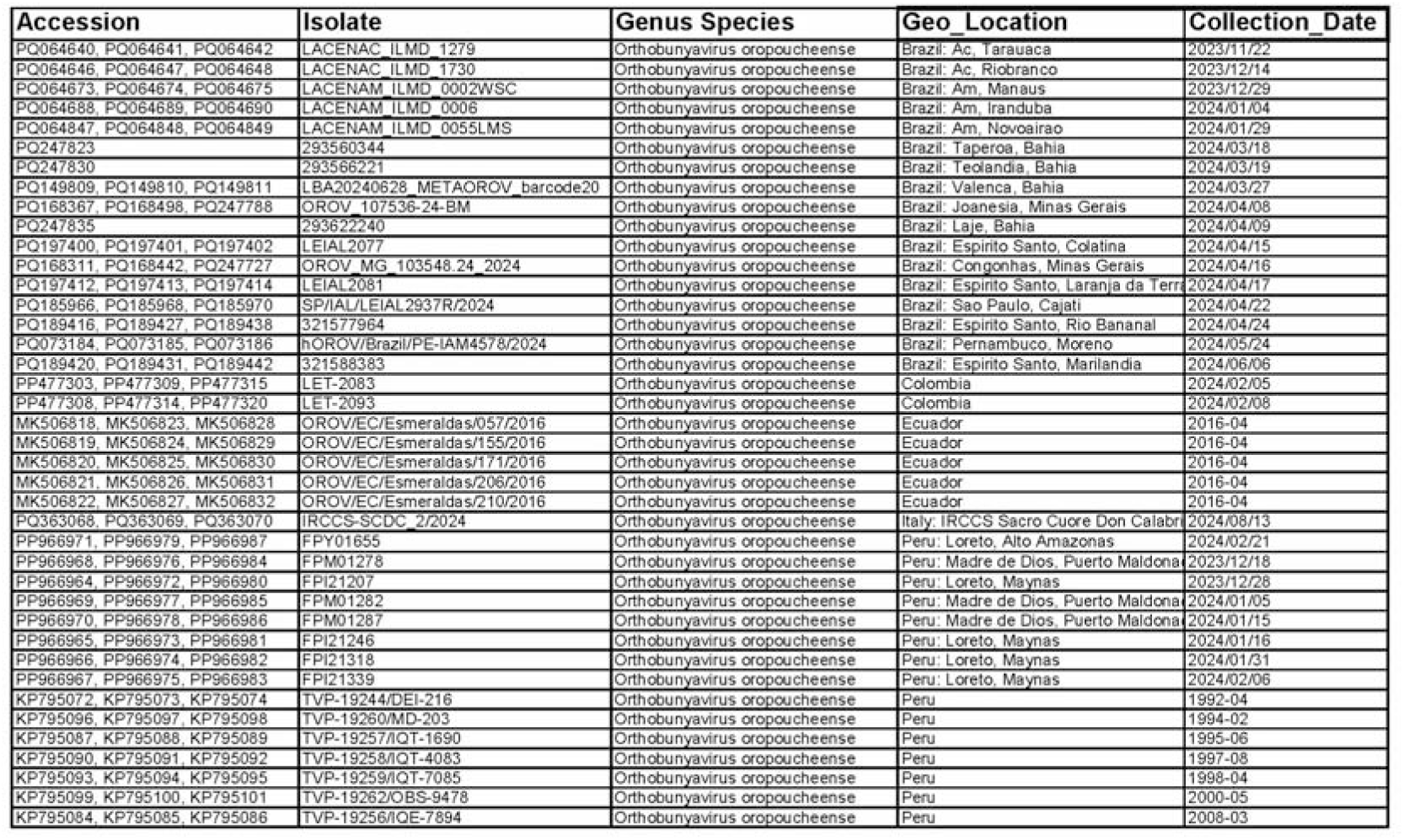
The accession number, isolate code, geographic location or origin, and collection date of strains included in phylogenetic analysis. All strains were human in origin.

## References

1. Wesselmann KM, Postigo-Hidalgo I, Pezzi L, de Oliveira-Filho EF, Fischer C, de Lamballerie X, et al. Emergence of Oropouche fever in Latin America: a narrative review. Lancet Infect Dis. 2024 Jan 25.

2. Sakkas H, Bozidis P, Franks A, Papadopoulou C. Oropouche Fever: A Review. Viruses. 2018 Apr 4;10(4).

3. Figueiredo LT. Emergent arboviruses in Brazil. Rev Soc Bras Med Trop. 2007 Mar-Apr;40(2):224–9.

4. Baisley KJ, Watts DM, Munstermann LE, Wilson ML. Epidemiology of endemic Oropouche virus transmission in upper Amazonian Peru. Am J Trop Med Hyg. 1998 Nov;59(5):710–6.

5. Aguilar PV, Barrett AD, Saeed MF, Watts DM, Russell K, Guevara C, et al. Iquitos virus: a novel reassortant Orthobunyavirus associated with human illness in Peru. PLoS neglected tropical diseases. 2011 Sep;5(9):e1315.

6. Watts DM, Russell KL, Wooster MT, Sharp TW, Morrison AC, Kochel TJ, et al. Etiologies of Acute Undifferentiated Febrile Illnesses in and near Iquitos from 1993 to 1999 in the Amazon River Basin of Peru. Am J Trop Med Hyg. 2022 Nov 14;107(5):1114–28.

7. Watts DM, Phillips I, Callahan JD, Griebenow W, Hyams KC, Hayes CG. Oropouche virus transmission in the Amazon River basin of Peru. Am J Trop Med Hyg. 1997 Feb;56(2):148–52.

8. Ladner JT, Savji N, Lofts L, Travassos da Rosa A, Wiley MR, Gestole MC, et al. Genomic and phylogenetic characterization of viruses included in the Manzanilla and Oropouche species complexes of the genus Orthobunyavirus, family Bunyaviridae. J Gen Virol. 2014 May;95(Pt 5):1055–66.

9. Navarro JC, Giambalvo D, Hernandez R, Auguste AJ, Tesh RB, Weaver SC, et al. Isolation of Madre de Dios Virus (Orthobunyavirus; Bunyaviridae), an Oropouche Virus Species Reassortant, from a Monkey in Venezuela. Am J Trop Med Hyg. 2016 Aug 3;95(2):328–38.

10. Tilston-Lunel NL, Shi X, Elliott RM, Acrani GO. The Potential for Reassortment between Oropouche and Schmallenberg Orthobunyaviruses. Viruses. 2017 Aug 11;9(8).

11. Hontz RD, Guevara C, Halsey ES, Silvas J, Santiago FW, Widen SG, et al. Itaya virus, a Novel Orthobunyavirus Associated with Human Febrile Illness, Peru. Emerging infectious diseases. 2015 May;21(5):781–8.

12. Hang J, Forshey BM, Yang Y, Solorzano VF, Kuschner RA, Halsey ES, et al. Genomic characterization of group C Orthobunyavirus reference strains and recent South American clinical isolates. PloS one. 2014;9(3):e92114.

13. Pinheiro FP, Travassos da Rosa AP, Gomes ML, LeDuc JW, Hoch AL. Transmission of Oropouche virus from man to hamster by the midge Culicoides paraensis. Science. 1982 Mar 5;215(4537):1251–3.

14. Cardoso BF, Serra OP, Heinen LB, Zuchi N, Souza VC, Naveca FG, et al. Detection of Oropouche virus segment S in patients and inCulex quinquefasciatus in the state of Mato Grosso, Brazil. Mem Inst Oswaldo Cruz. 2015 Sep;110(6):745–54.

15. Hoch AL, Pinheiro FP, Roberts DR, Gomes ML. Laboratory transmission of Oropouche virus by Culex Quinquefasciatus Say. Bull Pan Am Health Organ. 1987;21(1):55–61.

16. McGregor BL, Connelly CR, Kenney JL. Infection, Dissemination, and Transmission Potential of North American Culex quinquefasciatus, Culex tarsalis, and Culicoides sonorensis for Oropouche Virus. Viruses. 2021 Feb 2;13(2).

17. Pecor JE, Jones J, Turell MJ, Fernandez R, Carbajal F, O’Guinn M, et al. Annotated checklist of the mosquito species encountered during arboviral studies in Iquitos, Peru (Diptera: Culicidae). J Am Mosq Control Assoc. 2000 Sep;16(3):210–8.

18. Donayre Lopez CaMV, R. Caracterizacion Morfologica de Los Instars de Ochlerotatus serratues a partir de progenies producido en en laboratorio, Iquitos-Peru. Iquitos: Universidad Nacional de La Amazonia Peruana; 2013.

19. Pinheiro FP, Travassos da Rosa AP, Travassos da Rosa JF, Bensabath G. An outbreak of Oropouche virus diease in the vicinity of Santarem, Para, Brazil. Tropenmed Parasitol. 1976 Jun;27(2):213–23.

20. Acacia de Sa Pinheiro A, Morrot A, Chakravarty S, Overstreet M, Bream JH, Irusta PM, et al. IL-4 induces a wide-spectrum intracellular signaling cascade in CD8+ T cells. J Leukoc Biol. 2007 Apr;81(4):1102–10.

21. Almeida GM, Souza JP, Mendes ND, Pontelli MC, Pinheiro NR, Nogueira GO, et al. Neural Infection by Oropouche Virus in Adult Human Brain Slices Induces an Inflammatory and Toxic Response. Front Neurosci. 2021;15:674576.

22. Lorenz C, Azevedo TS, Virginio F, Aguiar BS, Chiaravalloti-Neto F, Suesdek L. Impact of environmental factors on neglected emerging arboviral diseases. PLoS neglected tropical diseases. 2017 Sep;11(9):e0005959.

23. Azevedo RS, Nunes MR, Chiang JO, Bensabath G, Vasconcelos HB, Pinto AY, et al. Reemergence of Oropouche fever, northern Brazil. Emerging infectious diseases. 2007 Jun;13(6):912–5.

24. Vasconcelos HB, Azevedo RS, Casseb SM, Nunes-Neto JP, Chiang JO, Cantuaria PC, et al. Oropouche fever epidemic in Northern Brazil: epidemiology and molecular characterization of isolates. J Clin Virol. 2009 Feb;44(2):129–33.

25. Sciancalepore S, Schneider MC, Kim J, Galan DI, Riviere-Cinnamond A. Presence and Multi-Species Spatial Distribution of Oropouche Virus in Brazil within the One Health Framework. Trop Med Infect Dis. 2022 Jun 20;7(6).

26. Mohapatra RK, Mishra S, Satapathy P, Kandi V, Tuglo LS. Surging Oropouche virus (OROV) cases in the Americas: A public health challenge. New Microbes New Infect. 2024 Jun;59:101243.

27. Moutinho S. Little-known virus surging in Latin America may harm fetuses. Science. 2024 Jul 26;385(6707):355.

28. Castilletti C, Mori A, Matucci A, Ronzoni N, Van Duffel L, Rossini G, et al. Oropouche fever cases diagnosed in Italy in two epidemiologically non-related travellers from Cuba, late May to early June 2024. Euro Surveill. 2024 Jun;29(26).

29. Organization PAH. Public Health Risk Assessment related to Oropouche Virus (OROV) in the Region of the Americas. 2024.

30. Penataro Yori P, Paredes Olortegui M, Schiaffino F, Colston JM, Pinedo Vasquez T, Garcia Bardales PF, et al. Etiology of acute febrile illness in the peruvian amazon as determined by modular formatted quantitative PCR: a protocol for RIVERA, a health facility-based case-control study. BMC public health. 2023 Apr 11;23(1):674.

31. Forshey BM, Guevara C, Laguna-Torres VA, Cespedes M, Vargas J, Gianella A, et al. Arboviral etiologies of acute febrile illnesses in Western South America, 2000-2007. PLoS neglected tropical diseases. 2010 Aug 10;4(8):e787.

32. Rainey JJ, Siesel C, Guo X, Yi L, Zhang Y, Wu S, et al. Etiology of acute febrile illnesses in Southern China: Findings from a two-year sentinel surveillance project, 2017-2019. PloS one. 2022;17(6):e0270586.

33. Quick J, Grubaugh ND, Pullan ST, Claro IM, Smith AD, Gangavarapu K, et al. Multiplex PCR method for MinION and Illumina sequencing of Zika and other virus genomes directly from clinical samples. Nature protocols. 2017 Jun;12(6):1261–76.

34. Wise EL, Marquez S, Mellors J, Paz V, Atkinson B, Gutierrez B, et al. Oropouche virus cases identified in Ecuador using an optimised qRT-PCR informed by metagenomic sequencing. PLoS neglected tropical diseases. 2020 Jan;14(1):e0007897.

35. NCBI Virus. 2004-ed. Bethesda (MD): Naitonal Library of Medicine (US), NAtional Center for Biotechnology Information.

36. Darriba D, Posada D, Kozlov AM, Stamatakis A, Morel B, Flouri T. ModelTest-NG: A New and Scalable Tool for the Selection of DNA and Protein Evolutionary Models. Mol Biol Evol. 2020 Jan 1;37(1):291–4.

37. Letunic I, Bork P. Interactive Tree of Life (iTOL) v6: recent updates to the phylogenetic tree display and annotation tool. Nucleic acids research. 2024 Jul 5;52(W1):W78–W82.

38. Edgar RC. Muscle5: High-accuracy alignment ensembles enable unbiased assessments of sequence homology and phylogeny. Nature communications. 2022 Nov 15;13(1):6968.

39. Kearse M, Moir R, Wilson A, Stones-Havas S, Cheung M, Sturrock S, et al. Geneious Basic: an integrated and extendable desktop software platform for the organization and analysis of sequence data. Bioinformatics. 2012 Jun 15;28(12):1647–9.

40. Stamatakis A. RAxML version 8: a tool for phylogenetic analysis and post-analysis of large phylogenies. Bioinformatics. 2014 May 1;30(9):1312–3.

41. Hellert J, Aebischer A, Wernike K, Haouz A, Brocchi E, Reiche S, et al. Orthobunyavirus spike architecture and recognition by neutralizing antibodies. Nature communications. 2019 Feb 20;10(1):879.

42. Gutierrez B, Wise EL, Pullan ST, Logue CH, Bowden TA, Escalera-Zamudio M, et al. Evolutionary Dynamics of Oropouche Virus in South America. Journal of virology. 2020 Feb 14;94(5).

43. Hang J, Yang Y, Kuschner RA, Evangelista J, Astete H, Halsey ES, et al. Genome Sequence of Bellavista Virus, a Novel Orthobunyavirus Isolated from a Pool of Mosquitoes Captured near Iquitos, Peru. Genome announcements. 2016 Nov 10;4(6).

44. Treangen TJ, Schoeler G, Phillippy AM, Bergman NH, Turell MJ. Identification and Genomic Analysis of a Novel Group C Orthobunyavirus Isolated from a Mosquito Captured near Iquitos, Peru. PLoS neglected tropical diseases. 2016 Apr;10(4):e0004440.

45. Tilston-Lunel NL, Hughes J, Acrani GO, da Silva DE, Azevedo RS, Rodrigues SG, et al. Genetic analysis of members of the species Oropouche virus and identification of a novel M segment sequence. J Gen Virol. 2015 Jul;96(Pt 7):1636–50.

